# Conversational artificial intelligence HeAlth supporT in Atrial Fibrillation Self-Management (CHAT-AF-S): rationale and randomised controlled trial design

**DOI:** 10.64898/2026.03.02.26347433

**Authors:** Liliana Laranjo, Aileen Zeng, Edel O’Hagan, Ritu Trivedi, Rahul Sathiaraj, Stuart Thomas, Aravinda Thiagalingam, Pramesh Kovoor, Gopal Sivagangabalan, Eddy Kizana, Saurabh Kumar, Jens Kilian, Simone Marschner, Tim Shaw, Clara K Chow

## Abstract

**Introduction:** Atrial fibrillation (AF), a common arrhythmia, is associated with impaired quality of life (QoL) and increased stroke risk and mortality. Clinical guidelines recommend leveraging digital technologies to support patient education and AF self-management. Conversational artificial intelligence (AI) technologies may support patient engagement with self-management by enabling human-like conversations. This study aims to evaluate the effectiveness of a conversational AI intervention (Conversational HeAlth supporT in Atrial Fibrillation Self-Management - CHAT-AF-S) in improving QoL in patients with AF.

**Methods and analysis:** CHAT-AF-S is a 3-month randomised controlled trial with 1:1 allocation and embedded process evaluation. We will randomise 480 adults (18 years of age and older) with documented AF to the CHAT-AF-S intervention or usual care. Primary outcome is the Atrial Fibrillation Effect on QualiTy-of-life (AFEQT) overall score. We will follow intention-to-treat principles and data analysts will be blinded. Intervention participants will be invited to complete a user experience survey and take part in an interview to explore the feasibility, acceptability, perceived utility, and barriers and enablers to implementing the intervention. Qualitative data will be analysed thematically.

**Ethics and dissemination:** Ethics approval was obtained from the Western Sydney Local Health District Human Ethics Research Committee (2023/ETH00765). Written and informed consent will be obtained from all study participants before commencing any study procedures. Results will be disseminated via peer-reviewed publications and presentations at international conferences.

**Declaration of Interests:** All investigators report nil conflicts of interest.

**Data Availability:** The data that supports this project are available from the corresponding author upon reasonable request.

**Trial registration:** Australian New Zealand Clinical Trials Registry ACTRN12623000850673 https://anzctr.org.au/Trial/Registration/TrialReview.aspx?id=386249

## INTRODUCTION

Atrial Fibrillation (AF) is the most common arrhythmia globally and its prevalence is rapidly increasing^1, 2^. AF is associated with increased risk of stroke, heart failure, and mortality^2^. AF symptoms such as palpitations, fatigue, and dyspnoea can significantly impair daily functioning and reduce quality of life^2, 3^. Guidelines emphasise patient empowerment as key to successful AF management across the components of the AF-CARE framework (C: Comorbidity and risk factor management; A: Avoid stroke and thromboembolism; R: Reduce symptoms by rate and rhythm control; E: Evaluation and dynamic reassessment)^4^. Appropriate AF management is key to improving health outcomes and quality of life for patients with AF.

Patient-centred AF management, comprising patient education, self-care support, and shared decision-making, can be enhanced by technology, as indicated in recent guidelines^4^. Digital AF patient support programs have shown promising albeit mixed results^5^, with some trials showing improvements in AF knowledge^6–9^ and medication adherence^10–12^. AF quality of life was shown to improve in one trial (n=120)^13^ evaluating a non-AI relational agent app [adjusted mean difference of 4.5 (95% CI 0.6 to 8.3) on the overall score of the Atrial Fibrillation Effect on QualiTy-of-life (AFEQT) questionnaire, at 1-month]^13^. However, the same non-AI relational agent evaluated in two larger trials (n=243, n=270) with a longer follow-up (12 months) failed to improve medication adherence or AF quality of life^14, 15^. The low levels of personalisation and “stickiness” (i.e. engagement propensity) of existing interventions may have limited their effectiveness and longer-term impact.

Recent advances in Artificial Intelligence (AI) have enabled the development of voice-based conversational agents that simulate human conversation through speech recognition and natural language processing. Conversational AI can support chronic disease self-management by engaging patients in conversations about their health, but randomised controlled trials remain sparse^16, 17^, particularly in AF. Our pilot trial (n=103) was the first to evaluate a voice-based conversational AI intervention for AF self-management^18^. Although the trial was underpowered to detect between-group differences, it showed a significant quality of life improvement pre-post in the intervention group [7.1 (95% CI 2.9 to 11.3) on the AFEQT overall score] and good acceptability^18, 19^. Qualitative analysis of participant feedback on this conversational AI^19^ contributed to the development of an improved intervention, named Conversational HeAlth supporT in Atrial Fibrillation Self-Management (CHAT-AF-S).

CHAT-AF-S is a 3-month self-management program comprising automated phone calls (i.e., voice-based conversational AI) and text-messages to support AF self-management. The aim of this randomised controlled trial is to evaluate the effectiveness of CHAT-AF-S in improving AF quality of life.

## METHODS

### Study Design

This study is a multicenter, single-blind, 1:1 (intervention:control) randomised controlled trial (RCT) with a 3-month follow-up, aiming to evaluate the effect of the CHAT-AF-S intervention on AF-related quality of life, compared with usual care (Figure 1). The trial has been registered in the Australian and New Zealand Clinical Trials Registry (Registration number: ACTRN12623000850673) and the study protocol is reported following the SPIRIT-AI Checklist^20^ for RCT protocols (Supplement 1) and the mobile health (mHealth) evidence reporting and assessment (mERA) checklist^21^ (Supplement 2).

**Figure 1.**
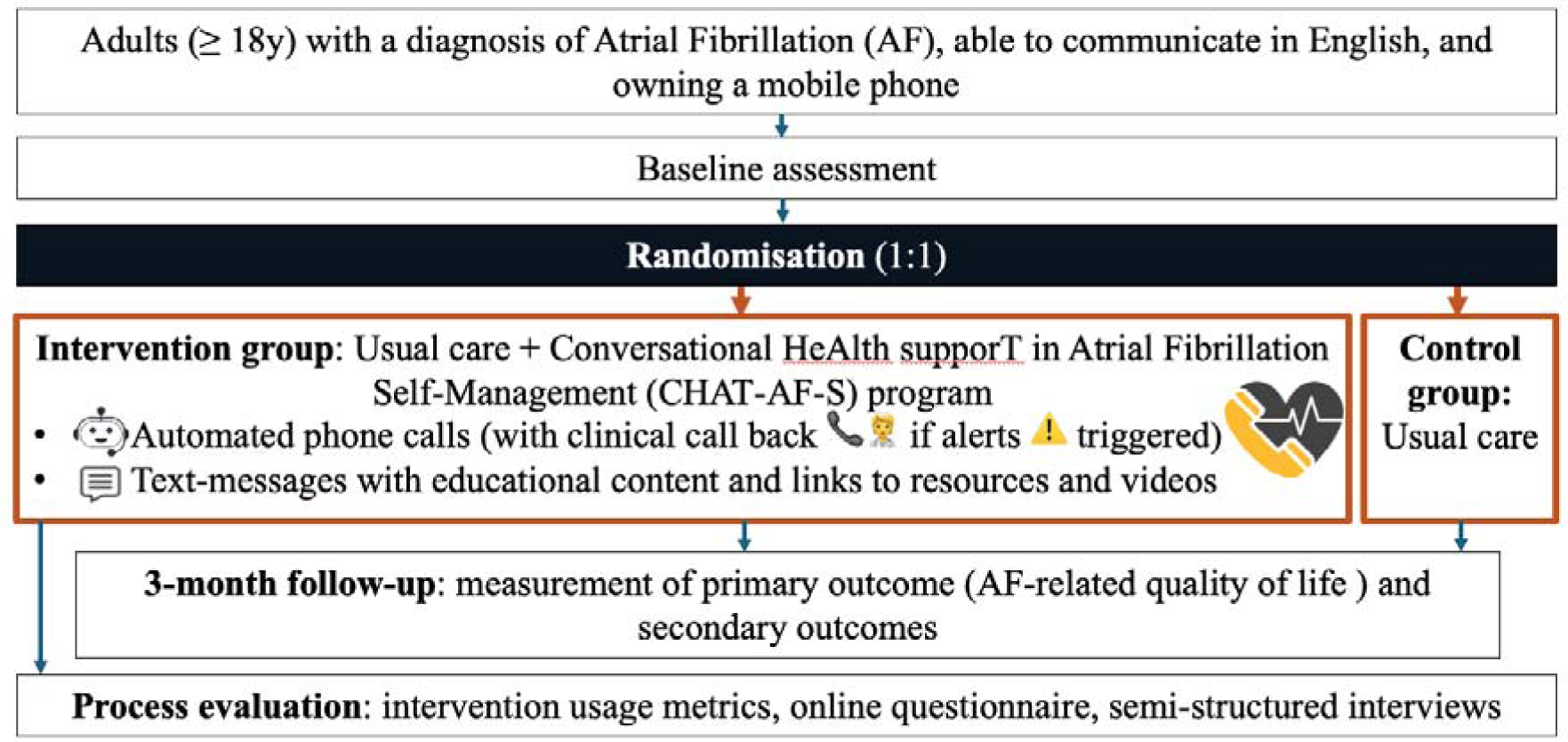
Study flow Diagram.

### Study Setting

This study is being conducted in Western and South Western Sydney, New South Wales, Australia. Participants will be recruited from cardiology inpatient and outpatient services in Westmead Hospital and Bankstown Hospital. The trial management team, located at the Westmead Applied Research Centre, is responsible for conducting and monitoring the trial across the two sites.

### Eligibility Criteria

Patients are eligible to participate if they (1) are 18 years of age or older, (2) have a documented diagnosis of AF, (3) have a mobile phone that is able to receive calls, (4) are able to receive text messages and open weblinks embedded in them, and (5) are competent with English, as ascertained by the study researcher. Participants will be excluded from the study if they (1) are on concurrent active interventional studies focused on AF education and self-management, (2) have a concomitant illness, physical impairment or mental condition which in the opinion of the study team/primary care physician could interfere with the conduct of the study including outcome assessment (e.g. hearing impairment not corrected by a digital hearing aid), (3) are pregnant, (4) have a medical illness limiting life expectancy, and (5) are unable or unwilling to provide written consent.

### Recruitment

Participants will be recruited from cardiology inpatient and outpatient services at Westmead Hospital and Bankstown Hospital, New South Wales, Australia. The research team and physicians will conduct weekly screening of upcoming outpatient appointments and inpatients ready for discharge to identify potentially eligible participants. The research team will approach identified patients either face-to-face during their visit or hospital admission, or over the telephone after their visit or hospital stay. The study will also be advertised through flyers in clinic waiting rooms and ward notice boards. The research team will invite eligible patients to participate in the study and provide them with a copy of the participant information and consent form. Participants will have the opportunity to ask questions before signing the consent form.

### Intervention

#### Design and development

Participants randomised to the intervention group will receive the CHAT-AF-S program in addition to their usual care; they will be provided with a letter to show their primary care doctor describing the AF self-management support that will be delivered by the CHAT-AF-S program, to support care coordination (Supplement 3). The “CHAT-AF-S” program was co-designed with clinicians (cardiology and primary care), academics (expertise comprising digital health, patient engagement, and behavioural science), and patients with AF, building on lessons learnt from our previous pilot trial evaluation, process evaluation and patient feedback (Australian New Zealand Clinical Trials Registry ACTRN12621000174886)^18, 19, 22^. The goal of CHAT-AF-S is to support patients with AF in managing their condition and thereby improve AF-related quality of life. The 3-month CHAT-AF-S program comprises six outreaches via automated phone calls (or text-message, after three unsuccessful call attempts at 24-hour intervals) and 30 educational text-messages containing AF-related information and self-management support, with links to online resources and videos (Figure 2). Calls and text-messages can be paused or discontinued based on participant request.

**Figure 2.**
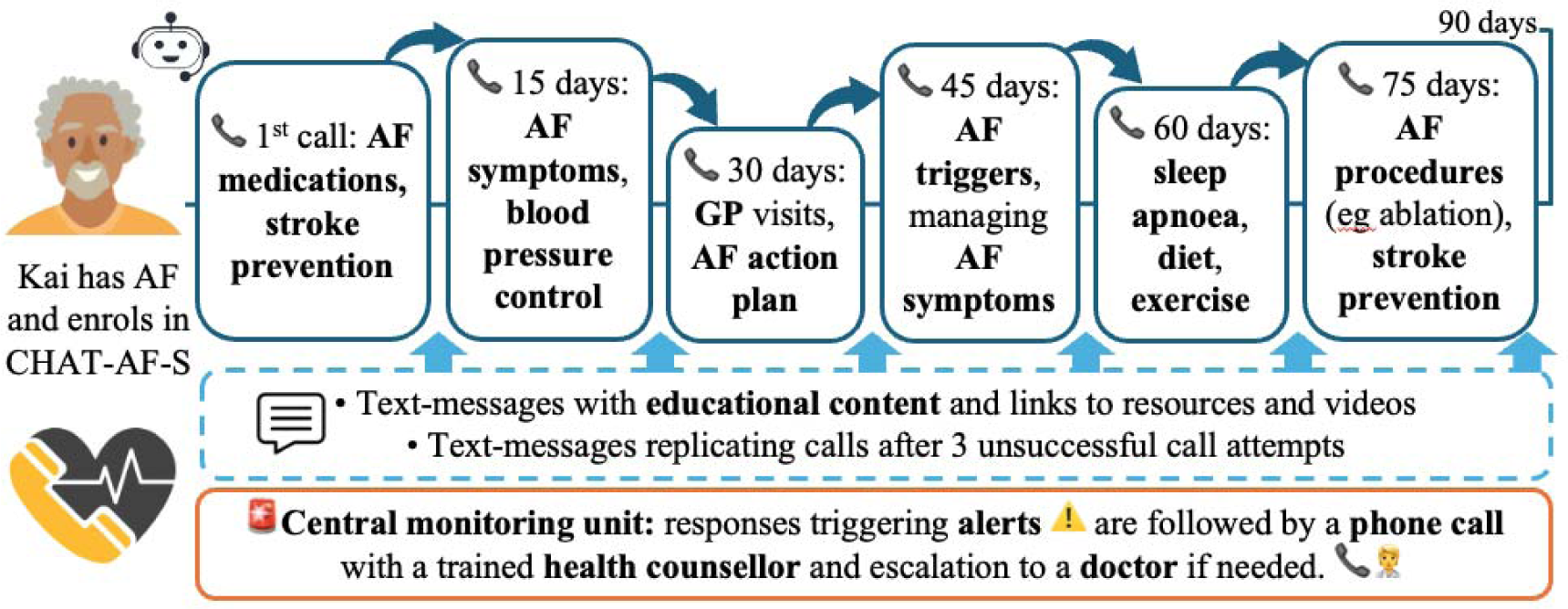
CHAT-AF-S Intervention.

The educational content and structure of the CHAT-AF-S program align with the AF-CARE Framework^4^ by providing tailored education on comorbidity and risk factor management, stroke prevention, symptom management, and rate and rhythm control^4^ (Figure 3). The program also enables regular assessment of risk and needs, monitoring of alerts generated by patient responses to risk assessment questions, and call back from health counsellor within two days after an alert is triggered, with clinical escalation as needed^4^. Behaviour change support in CHAT-AF-S focuses on lifestyle behaviours, medication adherence and management of risk factors and symptoms, and leverages behaviour change techniques aligning with the Behaviour Change Wheel^23, 24^ (Supplement 4).

**Figure 3.**
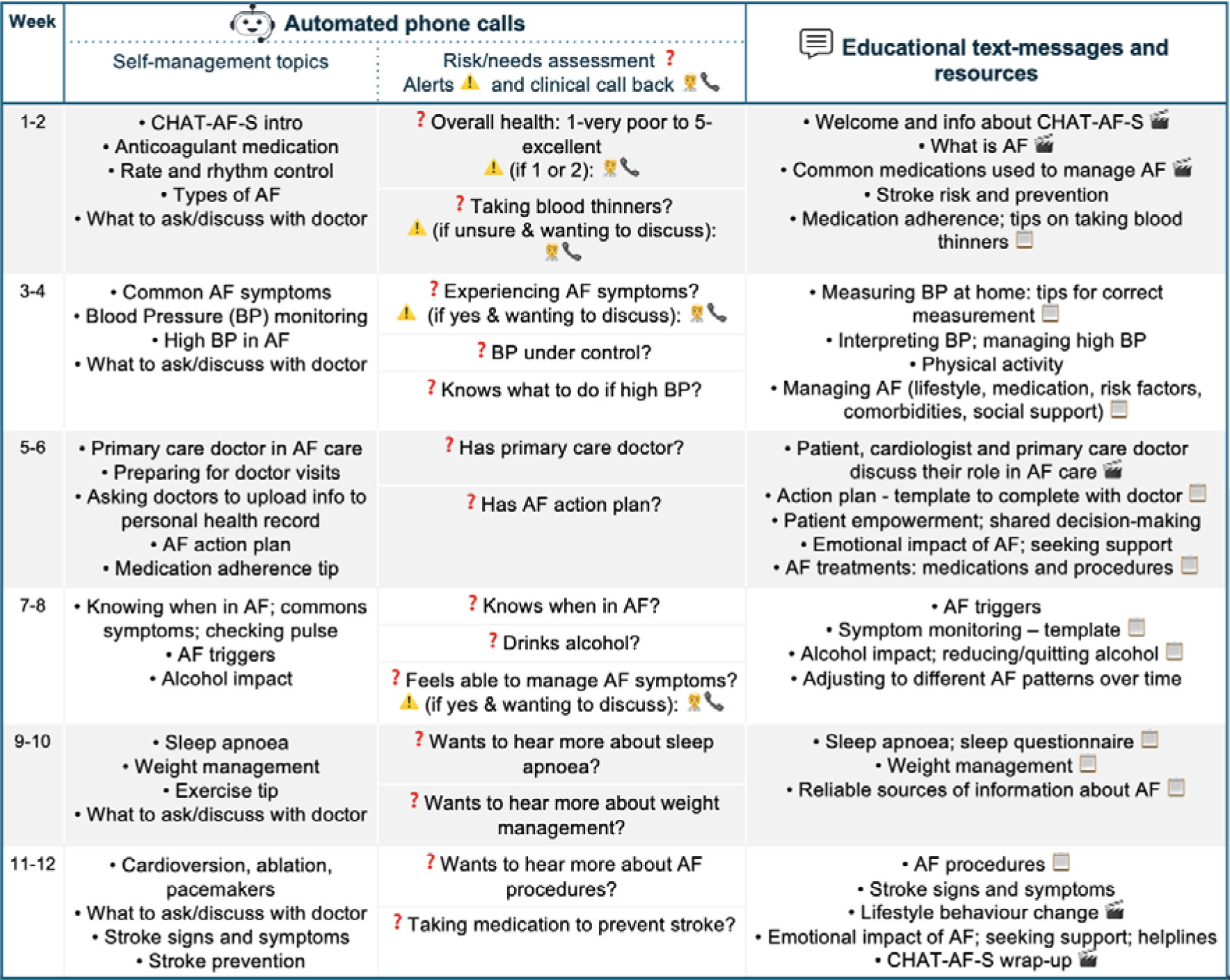
CHAT-AF-S structure and educational content to support AF self-management.

#### Automated phone calls

The automated phone calls in CHAT-AF-S are managed by a conversational AI system developed by Nuance^TM^. Design and development were done in the Nuance Mix conversational AI management platform (web-based cloud service), using Mix.nlu for the natural language understanding (identifying the semantic and syntactic elements from the user utterance to interpret intent and ‘understand’ what the user says), and Mix.dialog to design the conversational interface and dialog flows to engage the user. The Application is deployed on the Nuance Dialog as a Service (DLGaaS) platform and supports Automated Speech Recognition for Australian English. Reporting was implemented on the Nuance Insights platform connected to the Nuance DLGaaS instance. Additional platform capabilities for loading campaign lists and creating daily dialling campaigns for outbound calls are provided by Ericom^TM^ using Genesys.

The application is equipped with specific mechanisms for handling errors in recognition or understanding, including prompts paraphrasing the question and asking the user to repeat their answer, with the system moving on to the next step in the dialog flow after two attempts. Dual-Tone Multi-Frequency (DTMF) backup was also available for specific questions with numbered response options (eg call rating at the end of each call), allowing users to provide an answer by pressing keypad buttons when they encountered errors in recognition or understanding. To minimise interference from background noise in the flow of the conversation, speaking over prompts before they were complete is not supported.

#### Text messages

The CHAT-AF-S program includes two types of text-messages: call-replacement messages and scheduled educational messages. Call-replacement messages are sent to participants after three unsuccessful call attempts for one of the six outreaches. Replication of the structure and content of each of the six calls was developed in REDCap, matching the call scripts, including risk and needs assessment questions with alert responses triggering an email notification to the study team. Participants access the REDCap survey through a link included in the text-message sent by the study team.

The second type of messages are scheduled educational text-messages, including links to educational resources, webpages (e.g. Heart Foundation), questionnaires, templates, and videos. All educational resources are hosted on REDCap. Text-message delivery for call-replacement messages and scheduled educational messages will be done using TextCare and Digicuris platform (Westmead Applied Research Centre, University of Sydney).

### Control Group

The control group will receive usual care, which may include scheduled visits and clinical advice provided by their cardiologist or primary care doctor.

### Outcomes

The primary outcome of this trial is the Atrial Fibrillation Effect on QualiTy-of-life (AFEQT) overall score^25^ at 3 months between intervention and control groups. The AFEQT is reliable and validated 21-item questionnaire consisting of four domain scores (symptoms, daily activities, treatment concerns and treatment satisfaction) and an overall score. Scores range from 0 to 100 (higher scores associated with better quality of life). The overall AFEQT score is calculated using the symptoms, daily activity and treatment concern domain scores. A difference of 5 represents a clinically meaningful difference in AFEQT overall score^26–28^. Secondary outcomes include AFEQT domain subscale scores (symptoms, daily activities, treatment concerns, treatment satisfaction); pre-post AFEQT scores (overall and subscales) in the intervention group; AF knowledge measured with the Jessa Atrial Fibrillation Knowledge Questionnaire (JAKQ)^29^, a validated 16-item multiple choice questionnaire focusing on AF self-management and medications; Confidence in Atrial FibriLation Management (CALM)^30^, measured with the validated 16-item CALM questionnaire; treatment burden, measured with the validated 7-domain Treatment Burden Questionnaire (TBQ)^31, 32^; diet (measured with validated single-questions for fruit intake and vegetable intake)^33^; physical activity (measured using a validated single question^34^); alcohol intake; smoking; weight change from baseline; self-reported adherence to anticoagulant medication; and healthcare utilisation (primary care visits, cardiologist visits, emergency department visits, hospitalisations). All outcomes are measured at baseline and 3 months.

### Data collection and study procedures

Eligible individuals who have provided informed consent will be asked to complete a baseline questionnaire (Electronic Case Report Form; eCRF) to collect sociodemographic information. A medical history form will be completed by the research assistant by reviewing the electronic medical record. Participants in the intervention group will also complete a user experience survey to provide feedback on the intervention. Research Electronic Data Capture (REDCap) software^35^ will be used for data collection.

Engagement with the intervention will be assessed through intervention usage metrics. Participants in the intervention group will be invited to a semi-structured interview. We will use maximum variation sampling^36^ to interview a diverse sample of participants based on sociodemographic characteristics and their level of engagement with the intervention. The interview will be conducted via phone or online video call, audio-recorded, and transcribed verbatim. We will explore: patient experiences with CHAT-AF-S, including acceptability, engagement, and perceived utility; barriers and enablers to engagement; and impact of the intervention on the patient journey and care integration. Interviews will be conducted until conceptual depth^37^ is reached (we predict a minimum of 20 patients will be interviewed and estimate that each interview will last between 10 and 30 minutes).

### Randomisation and Blinding

Randomisation will be conducted electronically via the Research Electronic Data Capture (REDCap) software^35^. The software will automatically allocate participants to the intervention or control group, according to the randomisation sequence generated in R (using the randomiseR package) and uploaded to RedCap, ensuring allocation concealment. The software will randomise participants based on allocation ratio with stratification by sex and site; allocation concealment will be ensured as the randomisation sequence will not be accessible to the study team. Due to the nature of the study, participants and hospital-based cardiologists, as well as research study staff involved in the recruitment, data collection, and interviewing of participants, will not be blinded. However, the statistician analysing results will be blinded to the allocation.

### Sample size calculation

We estimated that a sample size of 480 would have 80% power to detect a difference of 7 in the overall score of the AFEQT questionnaire^25^ (α=0.05; Standard deviation=26), based on results from previous trials^38, 39^, accounting for 10% dropout. A difference of 5 represents a clinically meaningful difference in AFEQT overall score^26–28^.

### Data analysis

Statistical analysis will follow trial completion and database lock and will be conducted by a statistician blinded to group allocation, guided by a predefined statistical analysis plan. Analyses will follow the intention-to-treat principle (i.e., patients will be analysed within the group to which they were randomised). We will follow a published framework for intention-to-treat analysis that accounts for missing outcome data^40^: 1. Attempt to follow up all randomised participants, even if they choose to stop receiving the allocated treatment (i.e., discontinued treatment), as long as they have not withdrawn from the study; 2. Perform a main analysis of all observed data that are valid under a plausible assumption about the missing data; 3. Perform sensitivity analyses to explore the effect of departures from the assumption made in the main analysis; 4. Account for all randomised participants in the sensitivity analyses. For the primary and secondary continuous outcomes, groups will be compared at 3 months using Analysis of Covariance (ANCOVA), adjusted for corresponding baseline values. For dichotomous outcomes, groups will be compared using a log-binomial regression, while adjusting for corresponding baseline values as a fixed effect. Continuous variables will be reported in means and SDs, unless they are skewed in which medians and interquartile range will be reported, whereas categorical variables will be reported as frequencies and/or percentages. All continuous data will be checked for normality. Appropriate methods for analysing non-normally distributed data will be used where indicated. Effect estimates will be reported with 95% confidence intervals. All statistical tests will be two-tailed, and a significance level of 0.05 will be adopted throughout. All statistical analyses will be performed using R (version 4.1.0 or higher).

Data from interviews will be analysed using inductive thematic analysis^41^ of transcribed audio recordings. Results will be presented anonymously, ensuring that participants cannot be re-identified in any published results.

### Data management and monitoring

Data handling and storage will be conducted in accordance with the National Health and Medical Research Council guidelines and Australian Code for Responsible Conduct of Research. Identifiable data collected from this study will be stored on the secure research data store server provided by the University of Sydney and will only be accessible to study researchers. Questionnaire data will be collected electronically and stored on REDCap.

With participant consent, medical history data will be collected from the electronic medical record for the purpose of this study. As per the Health Records and Information Privacy Act 2002 No 71 (Schedule 1, Section 10 [1a]), the individual to whom the information relates will provide consent for use of their information, in line with the National Health and Medical Research Council’s National Statement. Data monitoring and trial audits will be conducted by an independent team with clinical trial expertise.

### Ethics and Dissemination

Ethics approval was obtained from the Western Sydney Local Health District Human Ethics Research Committee (2023/ETH00765; July 12, 2023). Written informed consent will be obtained from all study participants by a research assistant before commencing any study procedures, and a copy of the signed informed consent will be provided to each participant. Participation in the study is purely voluntary, and participants may withdraw at any time without consequences. The process of communicating information to participants and seeking their consent will ensure that a mutual understanding of the research and any associated expectations are clearly understood by participants.

Study results will be disseminated in peer-reviewed publications, scientific conferences, and to professional audiences and the public. The Open Science Framework will be used for sharing study materials (e.g., trial protocol, statistical analysis plan)^42^. A lay summary of the results will be prepared with our institution’s consumer group to support broader dissemination to interested community members.

## ANTICIPATED FINDINGS AND DISCUSSION

Recruitment started in November 2023, and as of December 2025 a total of 481 patients were recruited. Database lock and analysis are planned for early 2026. Results from this trial will provide evidence on the impact of CHAT-AF-S program in supporting patients with AF and improving their quality of life. The study will also provide insights into the acceptability, engagement, and perceived utility of AF self-management programs that leverage digital technologies such as conversational AI, with potential broader applicability in other chronic conditions. If successful, this AI-supported intervention could complement usual care for patients with AF, improving engagement in self-management and health outcomes.

## Data Availability

The data that supports this project are available from the corresponding author upon reasonable request.

## Acknowledgements

We acknowledge the nursing staff and doctors from Bankstown Hospital (Melissa Bryant), Westmead Hospital Cardiology Ward, Private Consulting Rooms, University Clinics and research officers (Kelly Chen, Diane Kancijanic, Carina Choy, Nayana Patel) for facilitating recruitment efforts. We would like to thank Dr Michelle Crockett for her input in the development of AF Educational content for our program.

## Author Contributions

LL drafted the initial draft, and all authors critically revised the manuscript, with LL, TS, CC taking a lead role. All authors approved the final version of the manuscript prior to submission.

