## Supplement for "Conversational artificial intelligence HeAlth supporT in Atrial Fibrillation Self-Management (CHAT-AF-S): rationale and randomised controlled trial design"

### Supplement 1. SPIRIT-AI Checklist - Recommended items to address in a protocol and related documents for clinical trials evaluating AI interventions^1^

| Section |  | SPIRIT AI Item | | Addressed on Page No ^b^ |
| --- | --- | --- | --- | --- |
| Title | 1 | Descriptive title identifying the study design, population, interventions, and, if applicable, trial acronym  Indicate that the intervention involves artificial intelligence / machine learning and specify the type of model.  Specify the intended use of the AI intervention. | | 1 |
|  |  |  |  | 1 |
| Trial registration | 2a | Trial identifier and registry name. If not yet registered, name of intended registry | | 3 |
|  | 2b | All items from the World Health Organization Trial Registration Data Set | | N/A |
| Protocol version | 3 | Date and version identifier | | N/A |
| Funding | 4 | Sources and types of financial, material, and other support | | 23 |
| Roles and responsibilities | 5a | Names, affiliations, and roles of protocol contributors | | 23 |
|  | 5b | Name and contact information for the trial sponsor |  | 23 |
|  | 5c | Role of study sponsor and funders, if any, in study design; collection, management, analysis, and interpretation of data; writing of the report; and the decision to submit the report for publication, including whether they will have ultimate authority over any of these activities |  | 23 |
|  | 5d | Composition, roles, and responsibilities of the coordinating centre, steering committee, endpoint adjudication committee, data management team, and other individuals or groups overseeing the trial, if applicable (see Item 21a for data monitoring committee) |  | 8 |
| Background and rationale | 6a | Description of research question and justification for undertaking the trial, including summary of relevant studies (published and unpublished) examining benefits and harms for each intervention  Explain the intended use of the AI intervention in the context of the clinical pathway, including its purpose and its intended users (e.g. healthcare professionals, patients, public).  Describe any pre-existing evidence for the AI intervention |  | 5-6 |
|  |  |  |  | 5-6 |
|  | 6b | Explanation for choice of comparators |  | 7 |
| Objectives | 7 | Specific objectives or hypotheses |  | 6 |
| Trial design | 8 | Description of trial design including type of trial (eg, parallel group, crossover, factorial, single group), allocation ratio, and framework (eg, superiority, equivalence, noninferiority, exploratory) |  | 7 |
| Study setting | 9 | Description of study settings (eg, community clinic, academic hospital) and list of countries where data will be collected. Reference to where list of study sites can be obtained  Describe the onsite and offsite requirements needed to integrate the AI intervention into the trial setting |  | 7 |
| Eligibility criteria | 10 | Inclusion and exclusion criteria for participants. If applicable, eligibility criteria for study centres and individuals who will perform the interventions (eg, surgeons, psychotherapists)  State the inclusion and exclusion criteria at the level of participants  State the inclusion and exclusion criteria at the level of the input data. |  | 8 |
|  |  |  |  | 8 |
| Interventions | 11a | Interventions for each group with sufficient detail to allow replication, including how and when they will be administered  State which version of the AI algorithm will be used.  Specify the procedure for acquiring and selecting the input data for the AI intervention.  Specify the procedure for assessing and handling poor quality or unavailable input data.  Specify whether there is human-AI interaction in the handling of the input data, and what level of expertise is required for users  Specify the output of the AI intervention  Explain the procedure for how the AI intervention’s output will contribute to decision-making or other elements of clinical practice. |  | 8-12 |
|  |  |  |  | 8-12 |
|  |  |  |  | 8-12 |
|  |  |  |  | 8-12 |
|  | 11b | Criteria for discontinuing or modifying allocated interventions for a given trial participant (eg, drug dose change in response to harms, participant request, or improving/worsening disease) |  | 8-12 |
|  | 11c | Strategies to improve adherence to intervention protocols, and any procedures for monitoring adherence (eg, drug tablet return, laboratory tests) |  | 8-12 |
|  | 11d | Relevant concomitant care and interventions that are permitted or prohibited during the trial |  | 8-12 |
| Outcomes | 12 | Primary, secondary, and other outcomes, including the specific measurement variable (eg, systolic blood pressure), analysis metric (eg, change from baseline, final value, time to event), method of aggregation (eg, median, proportion), and time point for each outcome. Explanation of the clinical relevance of chosen efficacy and harm outcomes is strongly recommended |  | 12-13 |
| Participant timeline | 13 | Time schedule of enrolment, interventions (including any run-ins and washouts), assessments, and visits for participants. A schematic diagram is highly recommended (see Figure) |  | 7 |
| Sample size | 14 | Estimated number of participants needed to achieve study objectives and how it was determined, including clinical and statistical assumptions supporting any sample size calculations |  | 14 |
| Recruitment | 15 | Strategies for achieving adequate participant enrolment to reach target sample size |  | 8 |
| Sequence generation | 16A | Method of generating the allocation sequence (eg, computer-generated random numbers), and list of any factors for stratification. To reduce predictability of a random sequence, details of any planned restriction (eg, blocking) should be provided in a separate document that is unavailable to those who enrol participants or assign interventions |  | 13-14 |
| Allocation concealment mechanism | 16b | Mechanism of implementing the allocation sequence (eg, central telephone; sequentially numbered, opaque, sealed envelopes), describing any steps to conceal the sequence until interventions are assigned |  | 13-14 |
| Implementation | 16c | Who will generate the allocation sequence, who will enrol participants, and who will assign participants to interventions |  | 13-14 |
| Blinding (masking) | 17a | Who will be blinded after assignment to interventions (eg, trial participants, care providers, outcome assessors, data analysts), and how |  | 13-14 |
|  | 17b | If blinded, circumstances under which unblinding is permissible, and procedure for revealing a participant’s allocated intervention during the trial |  | 13-14 |
| Data collection methods | 18a | Plans for assessment and collection of outcome, baseline, and other trial data, including any related processes to promote data quality (eg, duplicate measurements, training of assessors) and a description of study instruments (eg, questionnaires, laboratory tests) along with their reliability and validity, if known. Reference to where data collection forms can be found, if not in the protocol |  | 13 |
|  | 18b | Plans to promote participant retention and complete follow-up, including list of any outcome data to be collected for participants who discontinue or deviate from intervention protocols |  | 14 |
| Data management | 19 | Plans for data entry, coding, security, and storage, including any related processes to promote data quality (eg, double data entry; range checks for data values). Reference to where details of data management procedures can be found, if not in the protocol |  | 15 |
| Statistical methods | 20a | Statistical methods for analysing primary and secondary outcomes. Reference to where other details of the statistical analysis plan can be found, if not in the protocol |  | 14-15 |
|  | 20b | Methods for any additional analyses (eg, subgroup and adjusted analyses) |  | Will be reported in statistical analysis plan, which will be published in OSF before dataset unlock. |
|  | 20c | Definition of analysis population relating to protocol non-adherence (eg, as randomised analysis), and any statistical methods to handle missing data (eg, multiple imputation) |  |  |
| Data monitoring | 21a | Composition of data monitoring committee (DMC); summary of its role and reporting structure; statement of whether it is independent from the sponsor and competing interests; and reference to where further details about its charter can be found, if not in the protocol. Alternatively, an explanation of why a DMC is not needed |  | 15 |
|  | 21b | Description of any interim analyses and stopping guidelines, including who will have access to these interim results and make the final decision to terminate the trial |  | N/A |
| Harms | 22 | Plans for collecting, assessing, reporting, and managing solicited and spontaneously reported adverse events and other unintended effects of trial interventions or trial conduct  Specify any plans to identify and analyse performance errors. If there are no plans for this, explain why not. |  | Adverse events and serious adverse events will be recorded, with indication of whether they are likely related to the study or not. Performance errors of the AI intervention will be analysed as part of the process evaluation. |
| Auditing | 23 | Frequency and procedures for auditing trial conduct, if any, and whether the process will be independent from investigators and the sponsor |  | 15 |
| Research ethics approval | 24 | Plans for seeking research ethics committee/institutional review board (REC/IRB) approval |  | 15 |
| Protocol amendments | 25 | Plans for communicating important protocol modifications (eg, changes to eligibility criteria, outcomes, analyses) to relevant parties (eg, investigators, REC/IRBs, trial participants, trial registries, journals, regulators) |  | Protocol amendments will be approved by the Ethics Committee and updated in the trial registration. |
| Consent or ascent | 26a | Who will obtain informed consent or assent from potential trial participants or authorised surrogates, and how (see Item 32) |  | 15 |
|  | 26b | Additional consent provisions for collection and use of participant data and biological specimens in ancillary studies, if applicable |  | N/A |
| Confidentiality | 27 | How personal information about potential and enrolled participants will be collected, shared, and maintained in order to protect confidentiality before, during, and after the trial |  | 15 |
| Declaration of interests | 28 | Financial and other competing interests for principal investigators for the overall trial and each study site |  | 2 |
| Access to data | 29 | Statement of who will have access to the final trial dataset, and disclosure of contractual agreements that limit such access for investigators |  | 14 |
| Ancillary and post-trial care | 30 | Provisions, if any, for ancillary and post-trial care, and for compensation to those who suffer harm from trial participation  State whether and how the AI intervention and/or its code can be accessed, including any restrictions to access or re-use. |  | AI technology is proprietary and provided by Nuance. Intervention and study materials will be freely available in Open Science Framework. |
| Dissemination policy | 31a | Plans for investigators and sponsor to communicate trial results to participants, healthcare professionals, the public, and other relevant groups (eg, via publication, reporting in results databases, or other data sharing arrangements), including any publication restrictions |  | 15-16 |
|  | 31b | Authorship eligibility guidelines and any intended use of professional writers |  | We will follow ICMJE guidance |
|  | 31c | Plans, if any, for granting public access to the full protocol, participant-level dataset, and statistical code |  | 16 |
| Informed consent materials | 32 | Model consent form and other related documentation given to participants and authorised surrogates |  | 15 |
| Biological specimens | 33 | Plans for collection, laboratory evaluation, and storage of biological specimens for genetic or molecular analysis in the current trial and for future use in ancillary studies, if applicable |  | N/A |

^a^ It is strongly recommended that this checklist be read in conjunction with the SPIRIT 2013 Explanation & Elaboration for important clarification on the items; ^b^ Indicates page numbers to be completed by authors during protocol development.

Rivera SC, Liu X, Chan A-W, Denniston AK, Calvert MJ, Ashrafian H, et al. Guidelines for clinical trial protocols for interventions involving artificial intelligence: the SPIRIT-AI extension. The Lancet Digital Health 2020;2(10):e549-e560.

### Supplement 2. Mobile health (mHealth) evidence reporting and assessment (mERA) checklist^2^

| **Criteria** | **Item no** | **Notes** | **Section/Page** |
| --- | --- | --- | --- |
| **Infrastructure**  **(population level)** | 1 | Clearly presents the availability of infrastructure to support technology operations in the study location. This refers to physical infrastructure such as electricity, access to power, connectivity etc. in the local context. | 8-12 |
| **Technology platform** | 2 | Describes and provides justification for the technology architecture. This includes a description of software and hardware and details of any modifications made to publicly available software | 8-12 |
| **Interoperability/**  **Health information**  **systems (HIS) context** | 3 | Describes how mHealth intervention can integrate into existing health information systems. Refers to whether the potential of technical and structural integration into existing HIS or programme has been described irrespective of whether such integration has been achieved by the existing system | The platform is not integrated into existing health information systems. |
| **Intervention delivery** | 4 | The delivery of the mHealth intervention is clearly described. This should include frequency of mobile communication, mode of delivery of intervention (that is, SMS, face to face, interactive voice response), timing and duration over which delivery occurred | 8-12 |
| **Intervention content** | 5 | Details of the content of the intervention are described. Source and any modifications of the intervention content is described | 8-12 |
| **Usability/content**  **testing** | 6 | Describe formative research and/or content and/or usability testing with target group(s) clearly identified, as appropriate | 8-12 |
| **User feedback** | 7 | Describes user feedback about the intervention or user satisfaction with the intervention. User feedback could include user opinions about content or user interface, their perceptions about usability, access, connectivity, etc | 8-12 |
| **Access of individual**  **participants** | 8 | Mentions barriers or facilitators to the adoption of the intervention among study participants. Relates to individual-level structural, economic and social barriers or facilitators to access such as affordability, and other factors that may limit a user’s ability to adopt the intervention | 8-12 |
| **Cost assessment** | 9 | Presents basic costs assessment of the mHealth intervention from varying perspectives. This criterion broadly refers to the reporting of some cost considerations for the mHealth intervention in lieu of a full economic analysis. If a formal economic evaluation has been undertaken, it should be mentioned with appropriate references. Separate reporting criterion are available to guide economic reporting | To be conducted |
| **Adoption inputs/**  **programme entry** | 10 | Describes how people are informed about the programme including training, if relevant. Includes description of promotional activities and/or training required to implement the mHealth solution among the user population of interest | 8-12 |
| **Limitations for**  **delivery at scale** | 11 | Clearly presents mHealth solution limitations for delivery at scale | 8-12 |
| **Contextual**  **adaptability** | 12 | Describes the adaptation, or not, of the solution to a different language, different population or context. Any tailoring or modification of the intervention that resulted from pilot testing/usability assessment is described | 8-12 |
| **Replicability** | 13 | Detailed intervention to support replicability. Clearly presents the source code/screenshots/ flowcharts of the algorithms or examples of messages to support replicability of the mHealth solution in another setting | 8-12 |
| **Data security** | 14 | Describes the data security procedures/ confidentiality protocols | 8-12, 15 |
| **Compliance with**  **national guidelines**  **or regulatory statutes** | 15 | Mechanism used to assure that content or other guidance/information provided by the intervention is in alignment with existing national/regulatory guidelines and is described | 15-16 |
| **Fidelity of the**  **intervention** | 16 | Was the intervention delivered as planned? Describe the strategies employed to assess the fidelity of the intervention. This may include assessment of participant engagement, use of backend data to track message delivery and other technological challenges in the delivery of the intervention | To be conducted as part of process evaluation |

Agarwal S, LeFevre AE, Lee J, L’engle K, Mehl G, Sinha C, et al. Guidelines for reporting of health interventions using mobile phones: mobile health (mHealth) evidence reporting and assessment (mERA) checklist. bmj 2016;352.

### Supplement 3. Information letter for General Practitioner

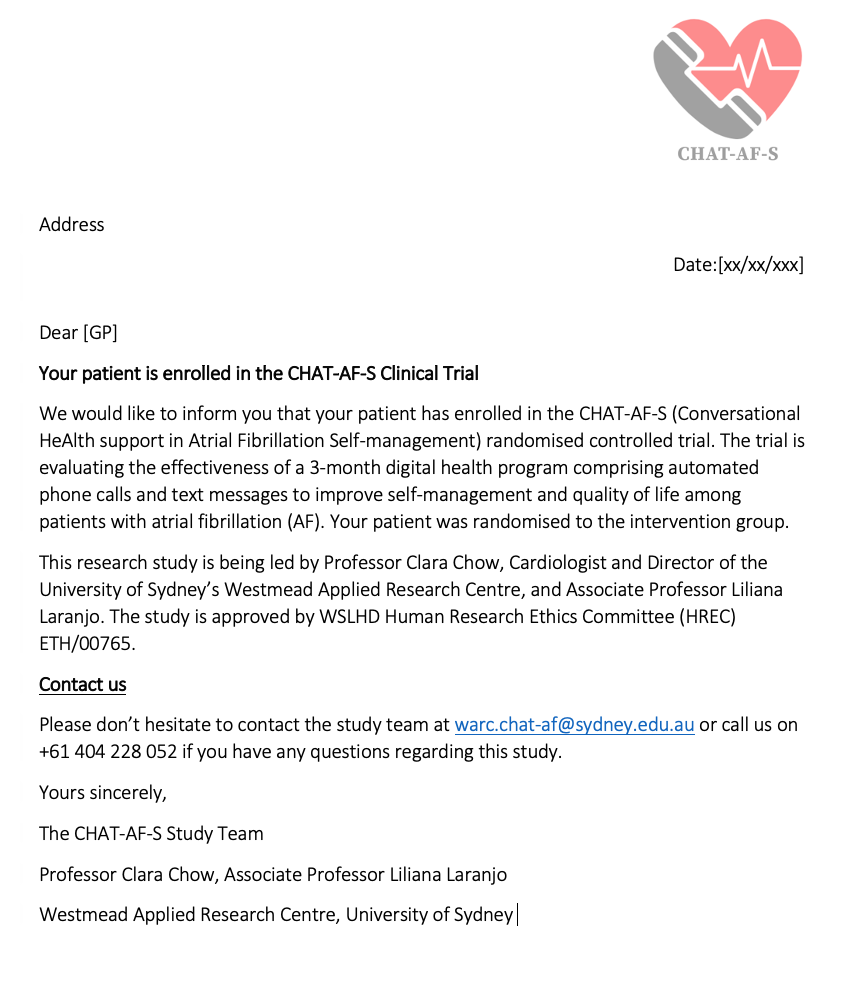

### Supplement 4. CHAT-AF-S behaviour change features

| **COM-B model component^3, 4^** | | **CHAT-AF-S features and BCTs** |
| --- | --- | --- |
| **Capability** | Psychological capability | Micro-learning through bite-sized education from credible source; information about health consequences and outcomes; action plan template; symptoms and triggers monitoring template; empowerment and shared decision-making support (eg “what to ask/discuss with doctor”) |
|  | Physical capability | Instruction on how to perform behaviours (eg measuring blood pressure) |
| **Opportunity** | Physical opportunity | Access to clinician escalation where needed, via call alerts and clinical call back |
|  | Social opportunity | Clinician reinforcement and support via call back |
| **Motivation** | Reflective motivation | Goal setting and action planning support; prompting discussions with doctors about preferences and goals |
|  | Automatic motivation | Suggestions for pairing behaviours (eg taking medication) with daily routines to form self-management habits; support for dealing with the emotional impact of AF |

Abbreviations: AF, Atrial Fibrillation; BCT, behaviour change technique; COM-B, Capability, Opportunity, Motivation, Behaviour model

**^3^**Michie S, Richardson M, Johnston M, Abraham C, Francis J, Hardeman W, et al. The behavior change technique taxonomy (v1) of 93 hierarchically clustered techniques: building an international consensus for the reporting of behavior change interventions. Annals of behavioral medicine 2013;46(1):81-95

**^4^**Michie S, Van Stralen MM, West R. The behaviour change wheel: a new method for characterising and designing behaviour change interventions. Implementation science 2011;6(1):42
